# EVALUATION OF THE PERFORMANCE OF THE ROUTINE EPIDEMIOLOGICAL SURVEILLANCE SYSTEM IN THE HEALTH DISTRICT OF TAMBACOUNDA (SENEGAL) IN 2020

**DOI:** 10.1101/2022.08.01.22278206

**Authors:** Tidiane Gadiaga, Mouhamadou Faly Ba, Siré Sagna, Bayal Cissé, Amadou Diallo, Samba Ndiaye, Boly Diop, Mbouna Ndiaye, Babacar Ndoye, Mamadou Sarifou Ba, Yoro Sall, Ndeye Mareme Sougou, Jean Augustin Diégane Tine, Mamadou Makhtar Mbacké Leye

## Abstract

**Introduction:** Epidemiological surveillance (ES) which is a continuous systematic process of data collection, analysis and interpretation for decision-making is of paramount importance for a good health system. Thus, to contribute to the improvement of the health system in Senegal, a study of the functioning of the routine epidemiological surveillance system was conducted in Tambacounda from S1 to S53 of the year 2020.

**Methodology:** A descriptive and analytical cross-sectional study was conducted from 1 to 17 July 2021. Comprehensive recruitment of the district’s 44 health care points was carried out. Data collection was carried out through a questionnaire prepared, pre-tested and administered to the 44 heads of public and non-public health facilities. The analyses were carried out with R software version 4.0.5.

**Results:** Of the 44 health facilities surveyed, 64% were public and 36% were non-public. The completeness and timeliness of the data were 100% and 97.5%, respectively. Suspected cases of tuberculosis were the most reported. For the providers surveyed (n=44), only 65.9% had knowledge of disease under epidemiological surveillance (DUES) and 93.2% managed suspected cases. On-site data analysis is only performed by 20.5% of providers. Only 38.6% of the service delivery point (SDP) had a health area card and the ES was under the responsibility of 77.3% of the paramedics. The training of ES officials was effective for only 45.5% of them. Despite the availability of dry tubes (69.8%), only 29.5% of PSD had COVID sampling equipment. The contribution of local authorities and technical and financial partners (TFP) to the SE was 22.7% and 29.5% respectively.

There was a statistically significant link between public SDP with knowledge of DUES (p <0.001), display of case definitions (p <0.001), feedback of reported cases, knowledge of indicators (p<0.001), existence of a health area map (p <0.001), advocacy with authorities (p=0.003), staff training (p=0.002), availability of DUES vaccines (p <0.001), availability of notification form (p<0.001) and partner contribution to ES activities.

**Conclusion:** Staff training, regular monitoring of ES activities with greater involvement of non-public structures, and the availability of inputs applied to the six pillars of the health system, are essential elements on which action must be taken. for an efficient ES system in the Tambacounda health district for the benefit of the country’s health system.

## Introduction

Epidemiological surveillance (ES) is a “continuous, systematic process of collecting, analyzing and interpreting data on specific health events, important for the planning, implementation and evaluation of public health practices, closely associated with their fair dissemination to those who need to be informed” [1]. It monitors the health system, which was defined by the World Health Organization (WHO) in its 2000 World Health Report, as the totality of organizations, institutions and resources devoted to the production of health interventions whose main objective is to improve, restore and preserve health [2]. This health system is based on six pillars: (i) health care service delivery, (ii) information - monitoring and evaluation - operational research, (iii) governance - leadership, (iv) human resources, (v) drug products - technology - vaccines and (vi) health financing [3,4]. In Senegal, the health system is pyramidal with three levels: (i) an operational level, the base of the pyramid consisting of health boxes, health posts, health centers and health districts, (ii) an intermediate level made by regional hospitals and the 14 medical regions, (iii) a central level, top of the pyramid with national hospitals, the various divisions and directorates of the Ministry of Health and Social Action [5]. At all levels of the health pyramid, routine ES is required and includes communicable diseases which are the most frequent causes of death and disability in the African region [6]. These priority diseases and events that are under SE are fifty-two [7] in addition to COVID-19 (integrated in 2020) and pose a significant threat to the well-being of African communities. In this regard, surveillance data will help guide health workers in the decisions to be taken for the implementation of appropriate control strategies and will guide prevention activities [6,8].” Improving the performance of the national health system requires in-depth and real-time knowledge of information on the occurrence or evolution of phenomena likely to affect public health [9].

However, the analysis of the different pillars that support health systems is not often applied to epidemiological surveillance. Also, surveillance data for communicable diseases may not be well recorded, reported and analyzed by different levels of the health system. As a result, opportunities for corrective action, appropriate public health responses and saving lives are lost.

Thus, it seems necessary to us to conduct a study to monitor more closely the functioning of the epidemiological surveillance system to contribute to the improvement of Senegal’s health system (in all its pillars) by strengthening the quality of epidemiological surveillance in the Health District (HD) OF Tambacounda.

Therefore, it will be specifically a question of measuring the completeness and timeliness of transmission of ES data, establishing the trend of priority diseases and COVID-19, describing the studied parameters of ES according to the different pillars of the Health System, analyzing the studied parameters of the ES system according to the type of health structures and determining the weak forces, opportunities and threats of the SE system of Tambacounda district in 2020.

## Data and Methods

### Study framework

The HD of Tambacounda is located in the department and the eponymous region. It is located in the south-east of Senegal with an area of 11,416 km^2^, an estimated population of 2020 to 295,018 inhabitants, a density of 25 inhabitants/km^2^ and a natural growth rate of 2.7% [10]. It consists of a public health institution level 2, a reference health center, 4 parapublic health centers, a garrison medical center, 24 public health posts, 5 private clinics, 2 paramedical practices, 4 corporate infirmaries, 2 private Catholic dispensaries. All health structures carry out routine epidemiological surveillance by respecting a circuit of transmission of information. The service delivery point (SDP) officers compile data from the various health facilities in their area of responsibility and then transmit them to the district malaria focal point. They also ensure the entry of this data every week in the DHIS2 platform [11].

### Type and period of study

This was a cross-sectional, descriptive, and analytical study of the functioning of the Epidemiological Surveillance system at the level of the HD of Tambacounda.

It covered the period from the first week (S1) to the fifty-third week (S53) of the year 2020. The data was collected in the period from 1 to 17 July 2021.

### Study population

These were the 44 SDP of the HD of Tambacounda.

### Inclusion criteria

Included in the study were all care benefit points of the HD of Tambacounda who performed ES during the 1st week to the 53rd week of the year 2020.

Epidemiological surveillance data from level 2 Public Health Facilities (PHF), private, parapublic and military structures are included in the reports of the “mother SDP” responsible for the health area.

### Non-inclusion criteria

Any SDP whose lead was not available to participate at the time of the study were not included in the study.

### Sampling procedure

For our work, an exhaustive recruitment of all the SPD of the HD of Tambacounda having carried out from the SE of S1 to S53 of the year 2020 was carried out.

### Data collection

#### Collection tool and data sources

The data were collected from a questionnaire consisting of six parts that integrated the six pillars of the health system.

For each of these parts, the questions concerned the elements of the epidemiological surveillance cycle, namely diagnosis and detection of cases, data collection and collection, data analysis and interpretation, information sharing, action, monitoring and evaluation of activities and training. The data source consisted of the DHIS2 platform to determine the completeness, timeliness and number of suspected cases of diseases under ES reported and the informants interviewed, i.e. SDP managers.

#### Collection method

Among the members of the district management team, two (2) investigators who had at least basic training in ES were selected and trained in one day on the administration of the questionnaire. They were accompanied by the expanded program on immunization (EPI)/ES Focal Point of the Tambacounda Medical Region. The questionnaire was tested at a health post near the Tambacounda health center. A restitution is organized on the same day with the members of the management team to share the difficulties and bring corrective measures.

After obtaining free and informed consent from SDP managers, the data was collected using the following techniques:

- Individual interviews, with the heads of structures to administer the questionnaire and collect their appreciation of the system in relation to its strengths, weaknesses, opportunities and threats (SWOT)
- Leveraging the DHIS2 platform to assess the completeness, promptness and trend of diseases

#### Operational definition of variables and indicators

- For the variables: these were:

**Public SDP:** These are the various health structures led by a civil servant, contractual of the Ministry, health development committees and / or local authorities.

**Non-public SDP**: These concerned private, confessional, military, paramilitary, parapublic (company infirmaries) health facilities.

- For the indicators: these were:

**Completeness**: Percentage of the number of reports received (numerator) over the number of reports expected (denominator), over a period

**Timeliness:** Percentage of the number of reports that were received on time (by the date and time set by the district in consultation with the providers) in the numerator over the number of reports expected in the denominator.

### Data analysis

#### Data entry

The collected data was entered into the Epi Info software version 7.2. Controls were integrated during the preparation of the input mask to limit input errors. A cleaning of the seized files had been done with the software’s analysis program and had corrected the outliers.

#### Methods of analysis

The analysis of the data was done in several stages. We first did a descriptive study that allowed quantitative variables to determine parameters (mean, standard deviation, median, extents, extremes,) and for qualitative variables (absolute, relative frequencies). Then, we then proceeded to an analytical study with bivariate analyses. To determine the existence of links between the different variables, we did an exact Fisher test with a significance threshold set at 0.05. The analyses were performed with R software version 4.0.5.

The Strengths, Weaknesses, Opportunities and Threats of the HD of Tambacounda in the context of the ES were identified by the informants.

##### Definition and choice of statistical units

These were all the SDP on which we expressed our results. They would also have escaped any previous studies of this kind and all met the above inclusion criteria. The choice thus fell on all the health structures that gravitated around the HD of Tambacounda, whatever the type.

#### Ethical considerations

Participation in this study was voluntary. It was done after free and informed consent. The evaluators administered a complete information form to the person in charge of the health structure. Confidentiality and anonymity were respected throughout the process. Apart from the completeness and promptness that were available in the national DHIS2 platform, the name of the SDP was hidden in the analysis of the other attributes of the ES.

## Results

### Descriptive part

At the end of our work, 44 health facilities were investigated. There were 64% of public health facilities, the rest (36%) were non-public SDP **(Figure 1)**.

**Figure 1:**
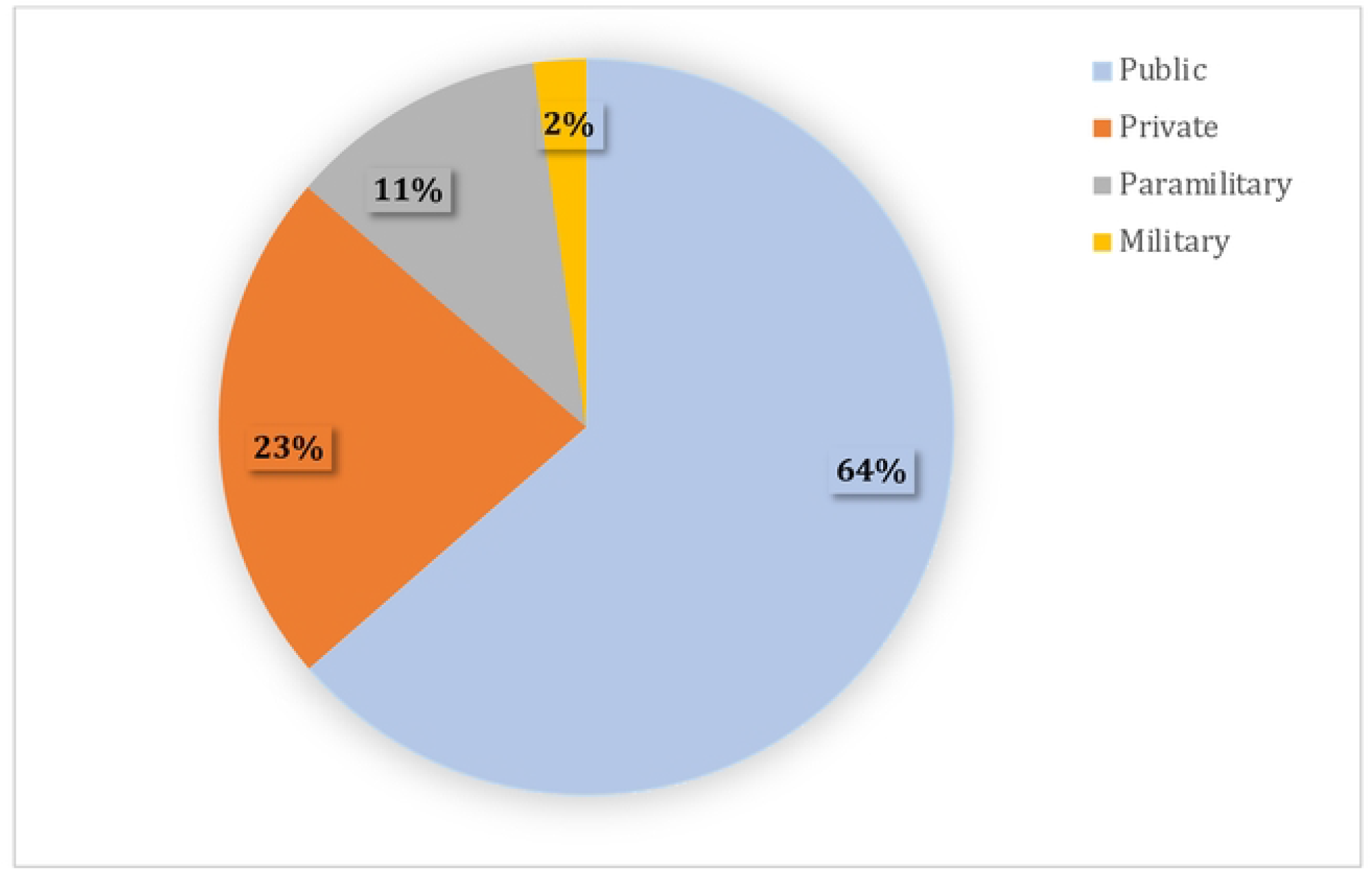
Distribution of the HD of Tambacounda structure types in 2020 (n=44)

#### Completeness

Of the 44 SDP in the district, 18 of them integrate their ES data into those of their parent health facility. Thus, for the completeness of the data, 26 health structures were monitored in the DHIS2 platform.

Completeness was 100% from the 1st to the 53rd week of the year 2020 for all 26 SDP in the district.

#### Promptitude

Of the 44 SDP in the district, 18 of them integrated their EM data with that of their parent health facility. Thus, for data completeness, 26 health facilities were monitored in the DHIS2 platform. Completeness was 100% from the 1st to the 53rd week of the year 2020 for all 26 SDP in the district.

#### Trend in SE and EPI-preventable diseases

Tuberculosis suspects were the most reported cases in the district in 2020. All other priority diseases under ES were also reported during the year at the district level **(Figure 2)**. Therefore, the district was not silent in any of the priority diseases under ES.

**Figure 2:**
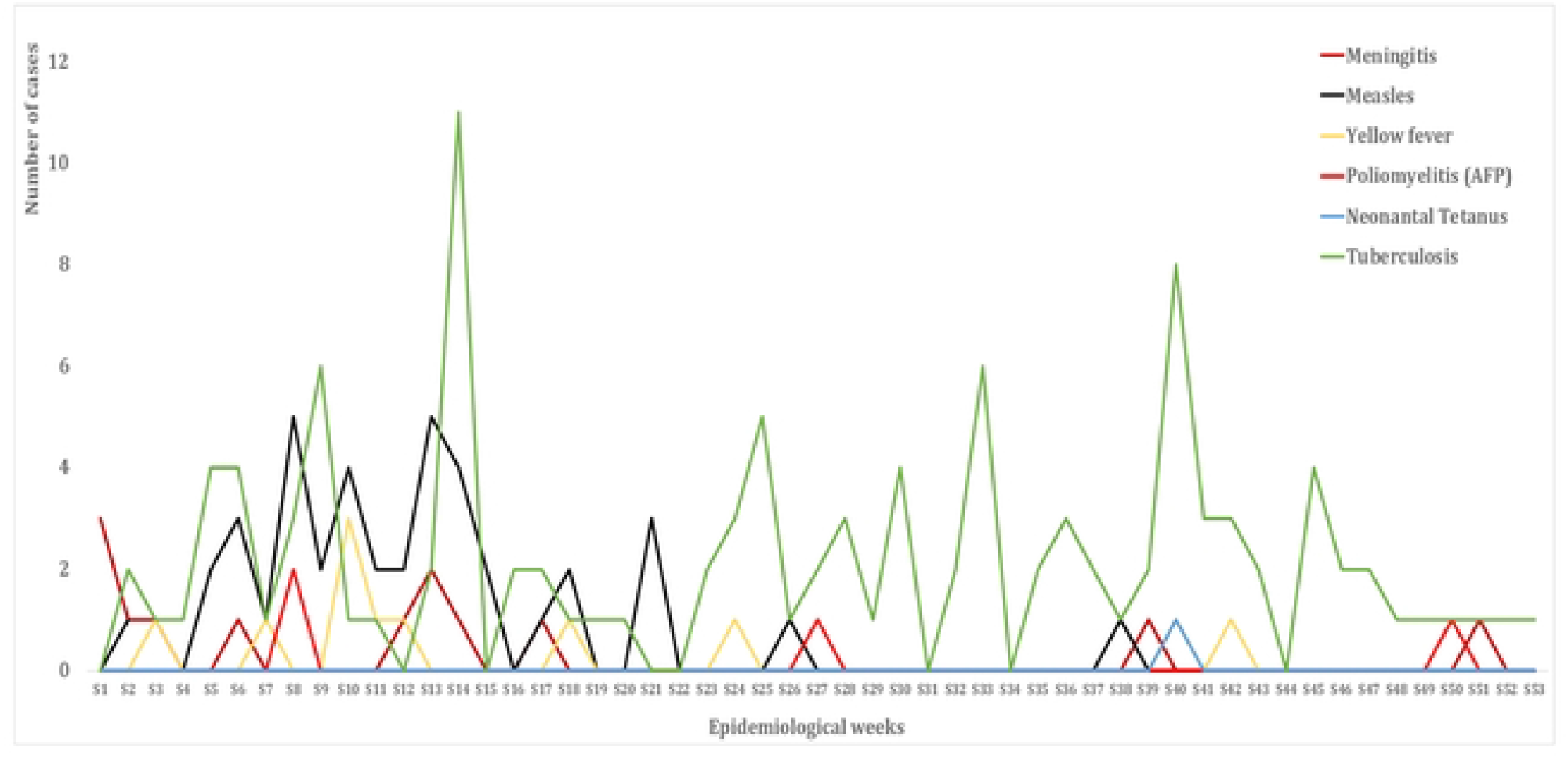
Evolution of suspected DUES cases preventable by vaccination from S1 to S53 of the year 2020 in the HD of Tambacounda

The district recorded the highest number of COVID-19 cases in the last eight weeks of 2020 with 89 positive cases out of 165 suspected cases between S45 and S53 of the year 2020 (**Figure 3)**. The peak of positive cases was reached at S47 with 18 cases.

**Figure 3:**
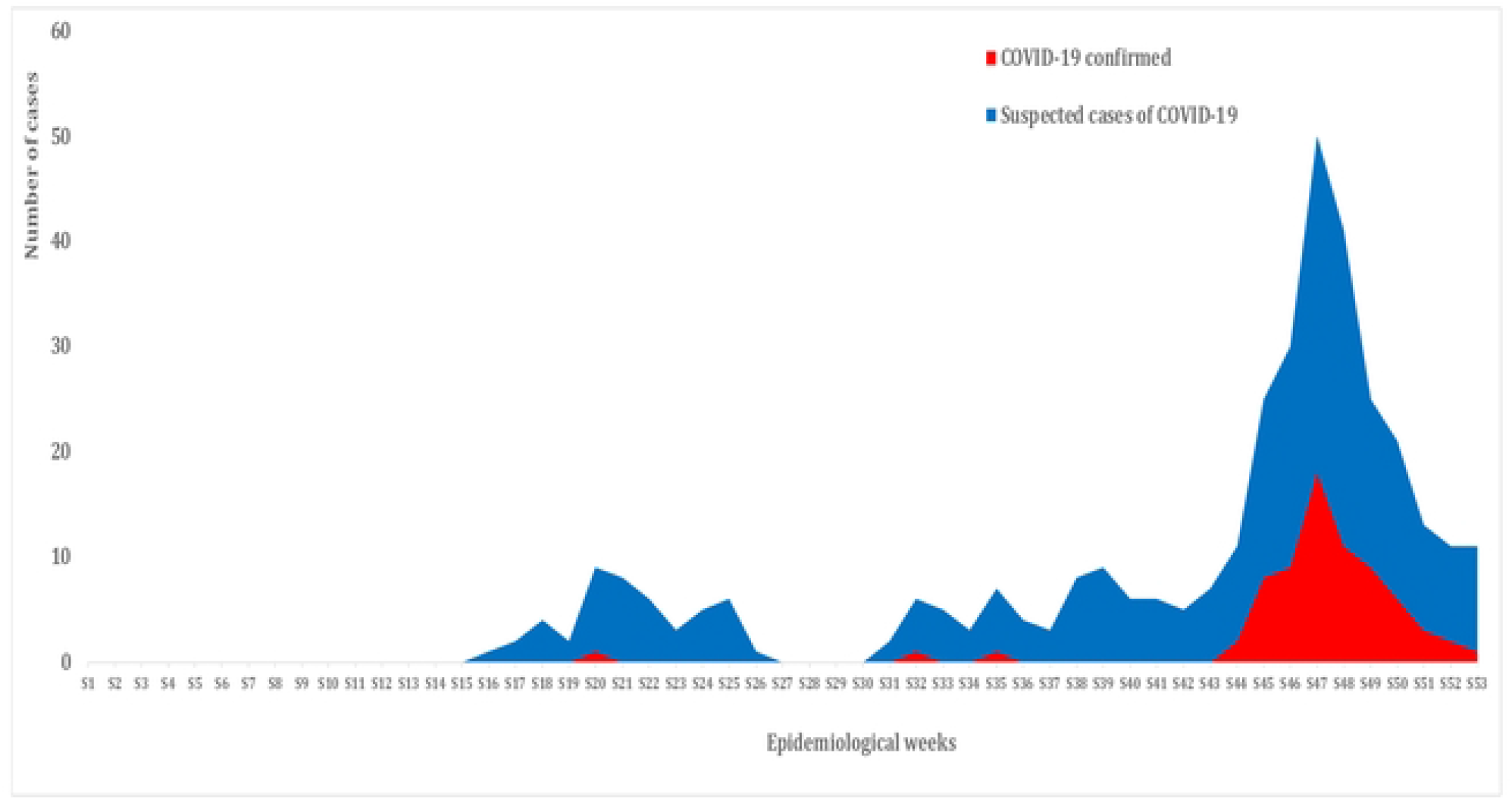
Evolution of suspected and confirmed cases of COVID-19 in 2020 at the HD of Tambacounda

#### Characteristics of SE according to the different pillars of the health system Provision of services

In our work, among the 44 SDP enrolled, 24 had an area of responsibility. The average number of polarized villages and/or neighborhoods was 20.2 with a standard deviation of 18.6. These 24 SDP covered on average 1 other health facility with a standard deviation of 1.8. The mean number of HVAC and functional health boxes were 4.7 (with a standard deviation of 4.0) and 2 (with a standard deviation of 1.6), respectively. The POL had an average of 2 traditional medicine practitioners (TMP) with a standard deviation of 1.9. However, of all the TMP identified in the district area, only 24% collaborate with SDP in the context of epidemiological surveillance.

Of all the SDP surveyed, only 65.9% had any knowledge of diseases and events under ES. But 93.2% of health facilities take care of suspected cases of DUES. However, at Community level the availability of ESM case definitions for a better understanding of ES diseases was low with 27.3% of cases (**Supplementary material 1**).

##### Information/Monitoring and Evaluation/Operational Research

Of the 44 SDP assessed, 65.9% recorded Immediately Notifiable Disease (IND) on notification forms. The collection and reporting of information at the SDP level was done on a weekly basis in 70.5% of cases; and that statement was made on Monday morning. In most cases the transmission of information was done through social networks (WhatsApp) with 54.5% of cases. However, compared to data analysis, only 20.5% of SDP analyzed their data on site and developed graphs of priority diseases. For operational research, only 11.4% of the district’s health facilities had to participate in ES research (**Supplementary material 2**).

Of the 24 SDP in the district with an area of responsibility, only 41.7% had to oversee the community level in the month before the survey date. Only 8% of health facilities had to carry out the 12 recommended monthly supervisions during the year.

Only 1 of the 44 SDP in the district (2.3%) received the recommended 12 monthly supervision missions from the ES FP district. A total of 11.4% of the SDP did not receive any oversight missions from the ES FP during 2020 **(Figure 4)**.

**Figure 4:**
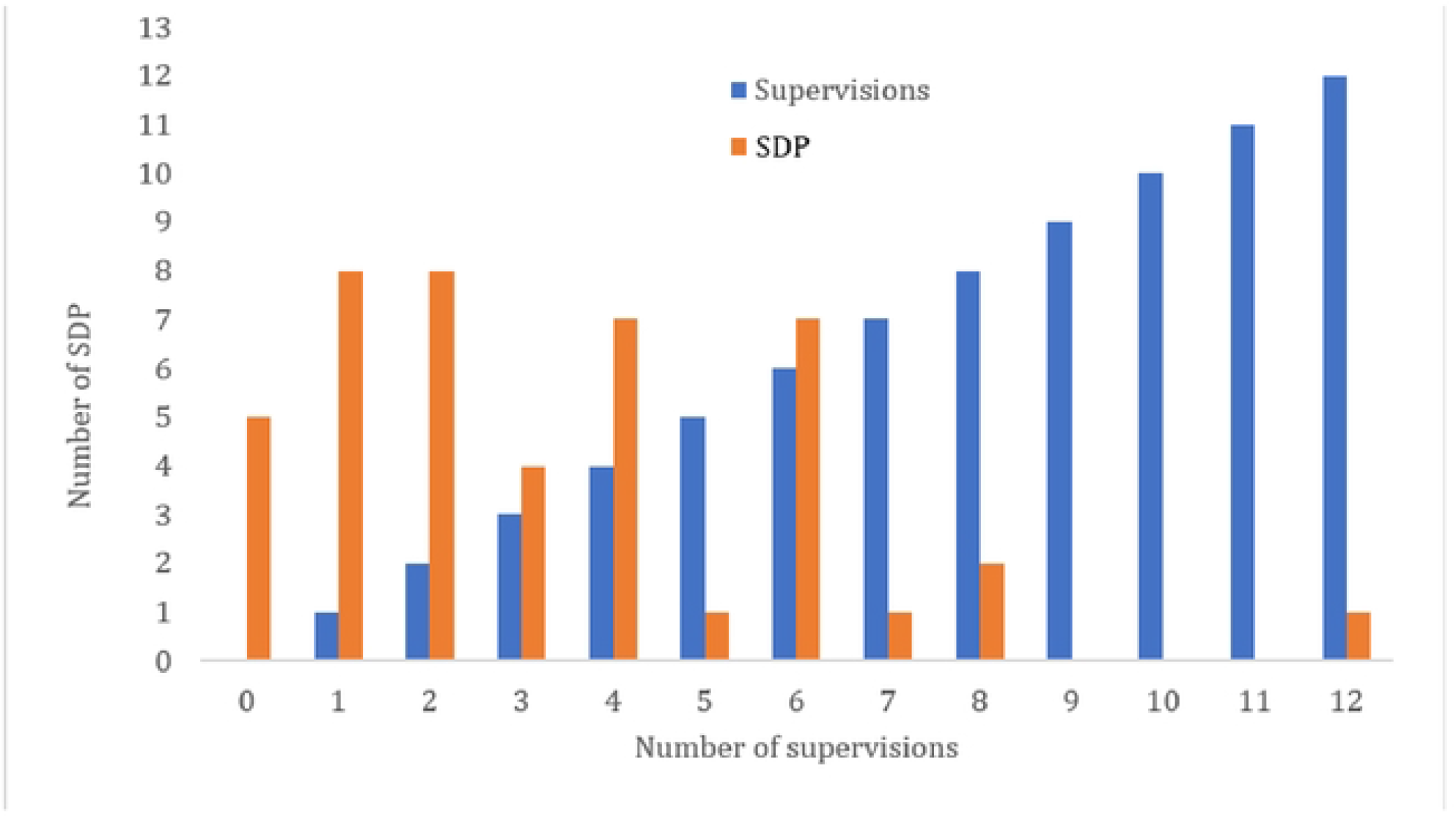
Distribution of the number of EPI/SE FP (Focal Point) supervisions in the district according to the SDP of the HD of Tambacounda in 2020 (n=44)

##### Governance and Leadership

The declaration of IND is the responsibility of the Head Nurses (HN) in 54.5% of cases. However, there is a standard method of reporting IND in 88.6% of the SDP surveyed. Only 25% of health managers have made a plea to the territorial authorities for their support to the ES (**Supplementary material 3**).

For a total of 44 SDP, 59% of them have never developed a Problem-Solving Plan (PSP) in 2020. No SDP was able to develop the 12 PRPs expected during the year

##### Human resources

Of the 44 SDP in our study, paramedics were responsible for ES in 77.3% of SDP. However, only 45.5% of ES managers have been trained in ES and/or epidemic management. The training of the rest of the staff was also almost non-existent because only 27.3% of the SDP had trained more than half of their staff in ES and/or epidemic management. No SDP (0%) involved more than half of its workforce in the ES. Only 11.4% of the SDP were able to supervise more than half of their workforce on the ES (**Supplementary material 4)**.

##### Drug products, technology, and vaccines

Appropriate ES inputs were only available in 9.1% of the district’s SDP. Despite the context of COVID-19, only 29.5% of SDP had COVID sampling equipment. However, there was a good availability of essential medicines for the management of suspected cases of DUES (81.8%). The same was true for the availability of vaccines against vaccines against vaccine-preventable DUES 56.8% of cases (**Supplementary material 5**).

##### Financing

There was a good contribution from SDP financial services to supervisory activities (95.5%). However, the contribution of local authorities and partners to ES activities was low at 22.7% and 29.5% respectively (**Supplementary material 6**).

The financial services of the SDP finance much more the transport of direct debits (95.2%).

Local and regional authorities are most often involved in the activities of the ES (70%) mainly in the purchase of medicines through the endowment funds allocated to health **(Figure 5)**.

**Figure 5:**
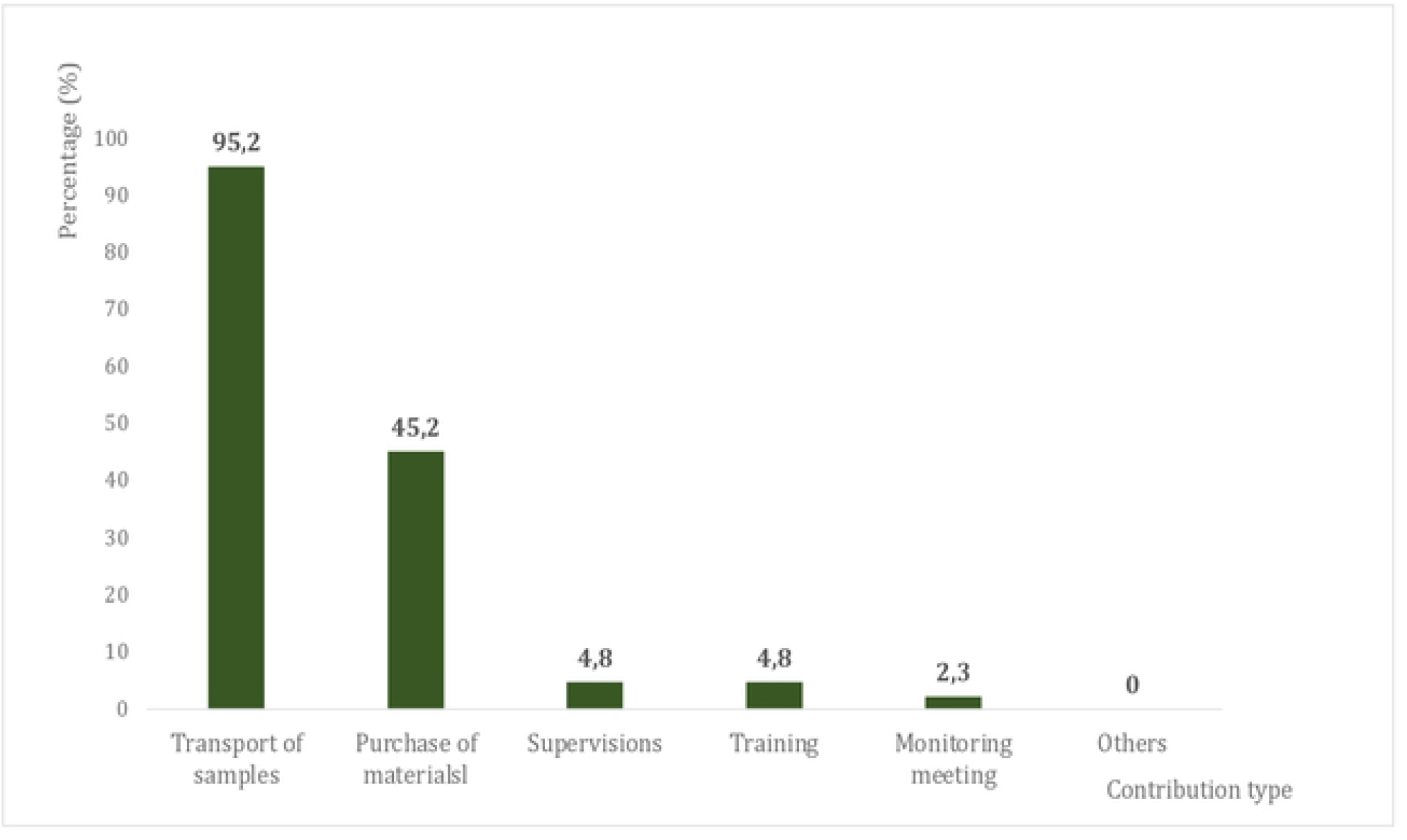
Contribution of Health Development Committee (HDC) and/or District SDP Financial Services to Surveillance Activities (n=42)

## Analytical part

Among those responsible for public SDP, 92.9% of them had knowledge of diseases under ES and among those in non-public SDP, only 18.8% of them had knowledge of diseases under ES. There was a statistically significant link between public SDP and knowledge of diseases under ES (p< 0.001). There was also a statistically significant link between public SDP and posting case definitions, feedback on reported and collected cases, analysis of data on the spot, existence of a health information circuit, advocacy with the authorities, training of the head of the ES structure, availability of vaccines, availability of essential medicines, contribution of local partners to ES activities **(Supplementary material 7)**.

The SWOT analysis of the ES system identified success and failure factors specific to the health system and other success and failure factors outside the health system **(Table I)**.

**Table I:**
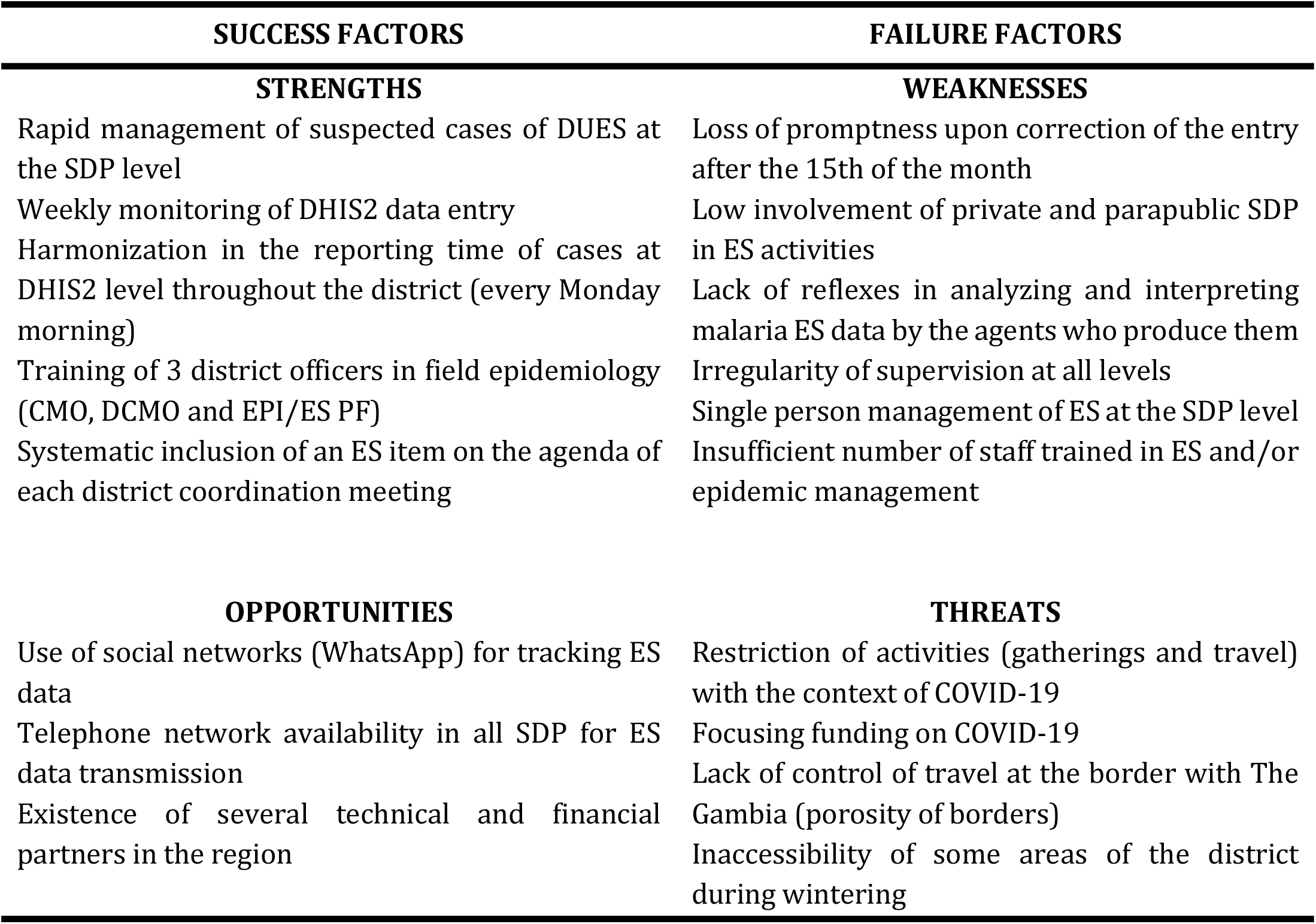
Analysis of strengths, weaknesses, opportunities and threats of the M&E system of the HD of Tambacounda, 2020

| SUCCESS FACTORS | FAILURE FACTORS |
| --- | --- |
| **STRENGTHS** | **WEAKNESSES** |
| Rapid management of suspected cases of DUES at the SDP level | Loss of promptness upon correction of the entry after the 15th of the month |
| Weekly monitoring of DHIS2 data entry | Low involvement of private and parapublic SDP in ES activities |
| Harmonization in the reporting time of cases at DHIS2 level throughout the district (every Monday morning) | Lack of reflexes in analyzing and interpreting malaria ES data by the agents who produce them |
| Training of 3 district officers in field epidemiology (CMO, DCMO and EPI/ES PF) | Irregularity of supervision at all levels |
| Systematic inclusion of an ES item on the agenda of each district coordination meeting | Single person management of ES at the SDP level |
|  | Insufficient number of staff trained in ES and/or epidemic management |
| **OPPORTUNITIES** | **THREATS** |
| Use of social networks (WhatsApp) for tracking ES data | Restriction of activities (gatherings and travel) with the context of COVID-19 |
| Telephone network availability in all SDP for ES data transmission | Focusing funding on COVID-19 |
| Existence of several technical and financial partners in the region | Lack of control of travel at the border with The Gambia (porosity of borders) |
|  | Inaccessibility of some areas of the district during wintering |

## Discussion

Our study aimed to assess the functioning of the system of routine epidemiological surveillance of diseases through the six pillars of the health system at the level of the health district of Tambacounda, operational level of the health pyramid of the health system of Senegal.

It was found that completeness and promptness were generally good, exceeding the national thresholds of 80%.” Regular monitoring of the level of ES data entry through district coordination bodies and exchange platforms (District WhatsApp Group) could explain this good performance. Keita had found the same performance in his study in Goudomp, Senegal in 2018 [12] On the other hand, Ly in his work in Madagascar in 2015, had found completeness that varied between 65% and 90% depending on the district [13].

### Provision of care services

The district was also successful in reporting all suspected cases of ES diseases. The mastery and application of case definitions by agents strongly contributed to this performance. This was contrary to Keita’s findings in his study in Goudomp where the district was silent for most diseases under ES [12]. However, in the context of service delivery, knowledge of DUES was much more pronounced among providers in the public sector (92.9%) than in the non-public sector (18.8%). The same was true for the availability of management tools essential for the proper delivery of ES services. The training and retraining of public sector employees to the detriment of the private sector would mainly explain this situation.

### Information, Monitoring and Evaluation and Operational Research

There was a harmonization of the frequency of reporting of suspected cases of DUES at the level of the different structures of the district; this thanks to the weekly reminders of the district surveillance focal point in the district information sharing group. The principle of reporting “Zero” cases is applied in most of the SDP in our study. Keita had found in his work, a performance of 71.4% in the PPS of Goudomp [12]. However, the analysis of surveillance data by the providers themselves was almost non-existent for both the public sector (32.1%) and the non-public sector (0.0%). Keita in his work at Goudomp had found similar results with only 29% of providers analyzing their surveillance data [12]. In the initial training of agents, no particular emphasis is placed on the analysis and interpretation of ES data. The same applies to the job descriptions of the officials responsible for surveillance. This is also reflected in the knowledge of performance indicators and the participation of agents in operational research on ES which are relatively low. However, performance feedback is effective for public SDP at the district’s monthly coordination meetings.

### Governance and Leadership

The health area map was available in 60.7% of THE SDP with Area of Responsibility. This performance was 78.6% in Keita’s study [12]. The lack of geographical coordinates of the health boxes and the polygons of the posts makes it difficult to make the maps. The only maps available are hand-drawn and were not accurate. Regarding the public SDP, 67.9% of them had set up a circuit for information at the level of Community actors. This result was close to that of Keita in his study in Goudomp with 78.6% of cases [12]. For better community participation in epidemiological surveillance activities, only 42.9% of public SDP held community feedback meetings compared to 0.0% of non-public SDP. The lack of funding (for the transport of community actors) would mainly explain the irregularity of community meetings. In our work, the development of problem-solving plans (PSPs) was almost non-existent, as was advocacy with territorial authorities. The lack of ownership of health management by the authorities through the health development committees does not motivate the leaders of the PPS to make regular advocacy and the elaboration of PSPs.

### Human Resources

The supervisor was a trained health worker in all SDP in the district. However, the involvement of other staff in the ES was low in the district; no structure involved more than half of its staff in the ES. This very often raises the problem of the correct continuity of the ES service during the absence of the person in charge of the structure. Training for the rest of the staff was not effective in the SDP. This performance was mainly linked to a regular transfer of staff to the capital and other regions of the northern and central zone of the country. Since 2017, the district has not received financial support for training officers on the expanded program on immunization and epidemiological surveillance. This last reason has even had an impact on the capacity building of community actors in both sectors. Series of trainings were included in the district’s work plan but were postponed due to the context of COVID-19, where gatherings were prohibited. The formative supervision of staff on the ES was also irregular at all levels of the health pyramid. This would mainly be related to the limitation of travel in the country (with the context of COVID-19) and the orientation of most health activities towards the management of COVID-19 cases.

### Drug products, technology and vaccines

Dry tubes to ensure timely sampling were available in 82.1% of SDP among those in the public sector and 17.9% in SDP in the private sector. At the Goudomp health district, Keita had found the same proportions of availability of sampling kits in the SDP of the public with a performance of 80% [12]. In our study, the low availability performance of tubes and sampling kits at the non-public sector level is explained by the fact that in case of presence of suspected cases, the head of the health area travels with his equipment to ensure the sampling. Compared to COVID-19, only designated public structures should have sampling equipment, hence this low performance for the overall SDP. However, essential medicines and vaccines against vaccine-preventable DUES were well available in SDP with areas of responsibility. The availability of vaccines and essential medicines are monitored monthly in all health posts and health centers in the district with area of responsibility. This orientation is mainly to avoid a break in vaccines, consumables, and essential medicines [14]. The availability of DUES case notification cards is also good in public SDP, linked to an initial allocation of management tools to all SDP by the district at the beginning of each year.

### Funding

As part of the mission of the HDC and administrative and financial service (AFS), the continuity of services is paramount; thus, they contributed to epidemiological surveillance activities in almost all of the SDP in our study. This contribution was much more marked for the transport of samples and the purchase of equipment because the speed of transport of samples to the medical region and the central level is evaluated each quarter according to the types of DUES during the national ES reviews [15]. On the other hand, for local and regional authorities, their contribution to surveillance activities at the SDP level was low (22.7%) even though health is considered a competence transferred to them. Despite the diversity of technical and financial partners at the Tambacounda level, their contribution to monitoring activities was low at the SDP level. The region’s health priorities were much more focused on malaria and maternal and child health. As a result, only 29.5% of all health facilities in the district were supported by TFP in terms of surveillance. However, with the context of COVID-19, TFP had also focused their interventions on the fight against this pandemic to the detriment of other diseases under epidemiological surveillance.

However, our study has some limitations. First, it is not representative of all health districts in Senegal. However, it can be very useful in other contexts, since in Senegal the health districts have the same functioning of the epidemiological surveillance system. Secondly, it did not consider the community component.

Finally, this study could also present a desirability bias because even with the fact of informing, the participants’ responses can be biased to project a better picture of their performance.

## Conclusion

With the re-emergence of diseases with epidemic potential, epidemiological surveillance is becoming increasingly essential in a well-functioning health system. For this reason, WHO had recommended to countries in the African Region the implementation of a Regional Project to Strengthen Surveillance Capacity in Africa in order to strengthen countries’ capacity for ES with the possibility of early detection and adequate case management as soon as possible. To do this, we need a strong health system in all these pillars. To meet this WHO directive, an evaluation of the epidemiological surveillance system across the pillars of the health system is crucial, especially since 2020 was particularly marked by a COVID-19 pandemic. It is in this context that we conducted this study which showed good completeness (100%), good promptness (97.5%) and notification of suspected cases of diseases under ES. However, despite the notable progress of the surveillance system, the analysis of the different determinants of ES at the level of each pillar of the health system has shown shortcomings at the level of all six pillars of the health system but which are much more marked at the level of non-public health structures than public structures. In perspective, it would be interesting to conduct a qualitative assessment of epidemiological surveillance data in the district to refine the recommendations in this area.

## Data Availability

All underlying data is available at https://docs.google.com/spreadsheets/d/1ZDU2j5fRGeQsqKoICkJpsTQi1hEoqz5L/edit?usp=sharing&ouid=108145215357620626491&rtpof=true&sd=true

## Acknowledgement

The thanks go to the Head Nurse, major of the reference health center of Tambacounda, the investigators and members of the executive teams of district and the medical region of Tambacounda without forgetting the drivers who ensured the transport of the teams.

## Financing

This research was funded by the Ministry of Health and Social Action of Senegal.

## Author Contributions

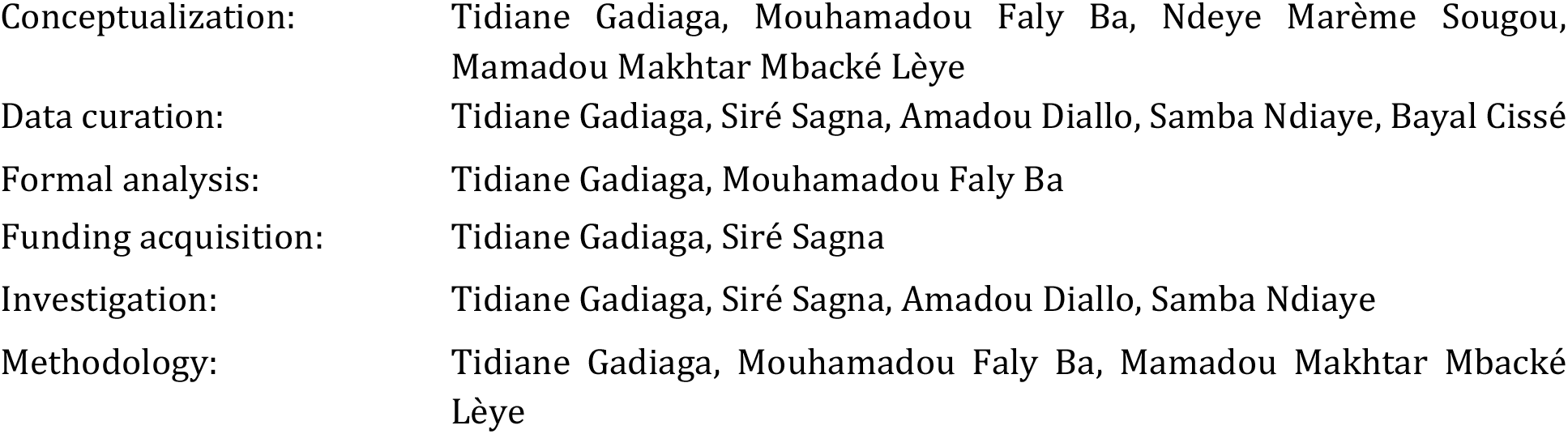
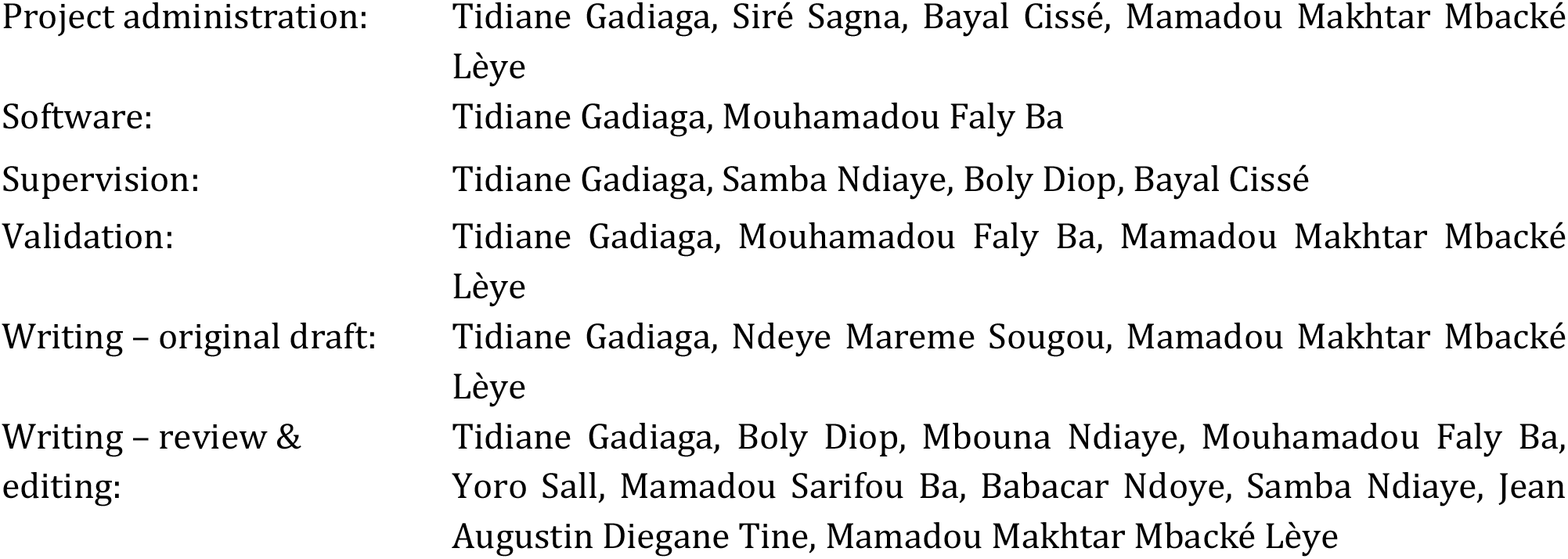

## Abbreviations

AFS: Administrative and Financial Service
CMAC: Community Monitoring and Alert Committee
CMO: Chief Medical Officer
DCMO: Deputy Chief Medical Officer
*DHIS2*: *District health information software*
DUES: Disease Under Epidemiological Surveillance
EPI: Expanded Program on Immunization
ES: Epidemiological surveillance
ES FP: Epidemiological Surveillance Focal Point
HCP: Home Care Provider
HD: Health District
HDC: Health Development Committee
HN: Head Nurse
IND: Immediately Notifiable Disease
MoHSA: Ministry of Health and Social Action
PHF: Public Health Facilities
PSP: Problem-Solving Plan
SDP: Service Delivery Point
SM: State midwife
TFP: Technical and financial partner
TMP: Traditional Medicine Practitioners
WHO: World Health Organization

## Supplementary materials

Supplementary material 1

Supplementary material 2

Supplementary material 3

Supplementary material 4

Supplementary material 5

Supplementary material 6

Supplementary material 7

